# Factors Associated with Stunting, Underweight, and Wasting Among Children Under Five in Bangladesh: Evidence from the MICS 2025 Survey

**DOI:** 10.64898/2026.10.21.26366175

**Authors:** Yesar Ahmed Oshan, Kakoli Khatun

**Author notes:** **Correspondence** Yesar Ahmed Oshan. **Funding information** No Funding.

## Abstract

**Background:** Childhood malnutrition remains a major public health concern in Bangladesh despite two decades of progress. Using the latest nationally representative data from Multiple Indicator Cluster Survey (MICS) 2025, this study assesses the prevalence of stunting, underweight, and wasting among children under five and examines their association with child, maternal, household, and regional characteristics.

**Methods:** We analyzed data on 21,736 children under five years of age from MICS 2025. Rao-Scott chi-squared tests and survey-weighted multivariable logistic regression were used to examine associations between socio-demographic factors and each malnutrition outcome.

**Results:** Weighted prevalence was 24% for stunting, 23% for underweight, and 13% for wasting. Stunting peaked at 12–35 months (AOR = 2.13, 95% CI: 1.82–2.50 for 12–23 months), whereas underweight was highest at 48–59 months (AOR = 1.88, 95% CI: 1.60–2.22). Girls had lower odds than boys of stunting (AOR = 0.92, 95% CI: 0.85–0.99), underweight (AOR = 0.93, 95% CI: 0.86–1.00), and wasting (AOR = 0.84, 95% CI: 0.76–0.92). Children from the richest households had substantially lower odds of stunting (AOR = 0.48, 95% CI: 0.41–0.56), underweight (AOR = 0.50, 95% CI: 0.42–0.58), and wasting (AOR = 0.71, 95% CI: 0.59–0.86). Maternal higher secondary education or above was protective against stunting (AOR = 0.57, 95% CI: 0.47–0.69) and underweight (AOR = 0.63, 95% CI: 0.51–0.76). Children in Sylhet had higher odds of stunting (AOR = 1.37, 95% CI: 1.15–1.64) and underweight (AOR = 1.40, 95% CI: 1.18–1.67) than those in Barishal.

**Conclusions:** Household wealth, children sex and birth order, maternal education, and geographic division are the main correlates of child malnutrition in Bangladesh. Equity-focused, region-specific interventions and continued national surveillance are needed to accelerate progress toward SDG 2.2.

## 1 INTRODUCTION

Child malnutrition is a major public health concern that includes both undernutrition, such as stunting, wasting, and being underweight, and overnutrition, such as overweight, obesity, and related diet-related diseases [1]. A child is considered malnourished when inadequate calorie or protein intake fails to meet the nutritional needs required for normal growth, when illness interferes with the body’s ability to utilize nutrients, or when energy intake significantly exceeds the body’s requirements [2].

The effects of inadequate nutrition during early childhood can persist throughout an individual’s life. Malnourished children are more vulnerable to infectious diseases, experience impaired cognitive and physical development, and face reduced productivity later in life, ultimately contributing to broader economic losses at the national level [3]. Malnutrition is estimated to contribute significantly to illness and mortality among children under five worldwide [4]. Conversely, rising rates of childhood overweight are contributing to an increasing risk of metabolic and cardiovascular diseases later in life [5]. As a result, countries such as Bangladesh are facing a “double burden” of malnutrition, in which undernutrition continues to coexist with growing levels of overweight and obesity [6].

According to the most recent Joint Child Malnutrition Estimates from UNICEF, the World Health Organization, and the World Bank, an estimated 150.2 million children under five were stunted globally in 2024 (23.2%), 42.8 million (6.6%) were affected by wasting (including 12.2 million severely wasted), and approximately 35.5 million were classified as overweight [7]. Although global stunting has fallen steadily since 1990, more than nine in ten stunted children still live in Asia and Africa, and current projections suggest the world will miss the 2030 stunting target by a wide margin [8]. In Bangladesh, an estimated 25% of children under five were stunted and 11% were affected by wasting based on the latest available national survey estimate [8].

Bangladesh continues to grapple with high levels of childhood stunting, surpassing the global average for children under five [9, 10]. A study drawing on data from 2007 to 2017 found that childhood stunting declined from 42% to 30% over that decade [11]. Similarly, data from the Multiple Indicator Cluster Survey (MICS) 2019 showed stunting falling from 42% in 2012 to 28% in 2019 [12]. However, the pace of decline remains steady rather than accelerating, raising concerns about the country’s ability to meet SDG target 2.2 [13].

Wasting remains a persistent global challenge, concentrated overwhelmingly in Asia and Africa (about three-quarters and one-fifth of cases, respectively) [14]. Recent global assessments, including the 2021 Global Nutrition Report, indicate that the world is not on track to achieve global wasting targets; progress toward both the 2025 World Health Assembly target of below 5% prevalence and the 2030 SDG-2 nutrition targets is further hindered by limitations in data availability and inconsistencies in cross-country comparability [15, 16, 17].

Bangladesh has made significant progress against undernutrition over the last two decades, yet the pace of improvement has slowed in recent years and appears to be reversing for some indicators. According to the Bangladesh Demographic and Health Survey 2022, stunting prevalence stood at 24% in 2022, down from 31% in 2017–18, while underweight prevalence remained largely unchanged at 22%; wasting, by contrast, rose from 8% to 11% over the same period [18]. Despite this body of evidence, no published study has yet examined the determinants of child malnutrition using Bangladesh’s most recent, nationally representative MICS7 (2025) data, leaving an important gap in up-to-date evidence to guide ongoing nutrition policy.

Malnutrition among children under five remains a multifaceted public health challenge in Bangladesh, driven by an interplay of household socioeconomic conditions and maternal and child healthcare practices. [19, 20, 21, 22]. Factors such as the child’s sex, place of residence, and maternal education were significantly associated with stunting among children under five years of age [23, 24, 19]. Sex, age and economic condition of the family appeared to be root causes of stunting [25, 26, 27, 28, 29, 30].

Malnutrition is a complex, multifactorial condition that can change over time, underscoring the need for continuous study. This research characterizes malnutrition using multiple indicators, assesses its prevalence, and investigates its association with key demographic, socioeconomic, environmental, and health-related factors. It also identifies the extent to which these factors contribute to stunting, wasting, and underweight among children in Bangladesh, using data from the latest Multiple Indicator Cluster Survey (MICS).

## 2 METHODS

For this study, the dataset was derived from the Bangladesh Multiple Indicator Cluster Survey (MICS7) 2025, which is a national-level survey conducted by the Bangladesh Bureau of Statistics (BBS) with the help of the United Nations Children’s Fund (UNICEF) [31]. The survey adopted a stratified two-stage sampling technique, whereby sampling strata consisted of urban and rural areas in each district. In the first stage, enumeration areas (EAs) were selected through probability proportional to size (PPS) sampling from each stratum. In the second stage, 20 households were systematically selected from each sample EA. Data collection for the survey took place from 24 February 2025 to 30 June 2025.

This study focused on three key indicators of childhood malnutrition: stunting, wasting, and underweight, each defined by anthropometric Z-scores based on the World Health Organization (WHO) 2006 Child Growth Standards. Stunting was identified by a height-for-age Z-score (HAZ) below −2 SD, wasting by a weight-for-height/length Z-score (WHZ) below −2 SD, and underweight by a weight-for-age Z-score (WAZ) below −2 SD. For each indicator, children were classified into one of two categories: those falling below the −2 SD threshold were considered affected, while those at or above it were not [32]. The initial dataset comprised 24,680 children under five years of age. After excluding 2,475 children with flagged (implausible) HAZ, WAZ, or WHZ values, 22,205 records remained, and a further complete-case exclusion of children with missing data on birth order, maternal age at birth, and mother’s age at first marriage yielded a final sample of 21,736 children under five years of age (weighted *N* = 21,699) who had complete information on all study variables.

A range of child, maternal, and household-level characteristics were included as explanatory variables in the analysis. These included the child’s age, sex, and birth order; maternal age at birth and at first marriage; and household-level factors such as place of residence, administrative division, household size, number of living children in the household, and the sex, education, and religion of the household head. Household wealth index and maternal education were also considered. The specific categories used to classify each covariate are presented in Table 1.

**TABLE 1.** Description of study covariates and their categories.

| Covariate | Description | Categories |
| --- | --- | --- |
| Child's Age (months) | Age of the child at the time of survey | <6, 6–11, 12–23, 24–35, 36–47, 48–59 |
| Sex | Sex of the child | Male, Female |
| Birth Order | Birth order of the child | 1<sup>st</sup>, 2<sup>nd</sup>–3<sup>rd</sup>, ≥ 4<sup>th</sup> |
| Maternal Age at Birth | Mother's age at the child's birth | <20, 20–34, ≥ 35 years |
| Residence | Place of residence | Urban, Rural |
| Division | Administrative division of residence | Barishal, Chattogram, Dhaka, Khulna, Mymensingh, Rajshahi, Rangpur, Sylhet |
| Household Size | Total number of household members | ≤ 3, 4–6, ≥ 7 |
| Number of Living Children | Total number of living children in the household | ≤ 2, ≥ 3 |
| Household Head's Sex | Sex of the household head | Male, Female |
| Religion | Household's religious affiliation | Islam, Other |
| Wealth Index | Household socioeconomic status | Poorest, Poorer, Middle, Richer, Richest |
| Maternal Education | Highest educational attainment of mother | Pre-primary or none, Primary, Secondary, Higher secondary or above |
| Household Head's Education | Highest educational attainment of household head | Pre-primary or none, Primary, Secondary, Higher secondary or above |
| Mother's Age at First Marriage | Mother's age during first marriage | <15, 15–20, ≥ 20 years |

Descriptive statistics were used to describe the study population and weighted prevalence estimates were calculated separately for stunting, underweight and wasting. First, we assessed bivariate associations between socio-demographic characteristics and each malnutrition outcome using a Rao–Scott second-order corrected chi-squared test [33]. Significant variables were included in the multivariate models. Separate survey-weighted logistic regression models [34], both univariate and multivariate, were fitted for each outcome, yielding crude and adjusted odds ratios (CORs and AORs) with corresponding 95% confidence intervals. We considered P-value less than 0.05 as statistically significant. All analyzes were done in R [35].

## 3 RESULTS

Weighted estimates showed that 24% of children were stunted, 23% were underweight and 13% were wasted (Figure 1). Chi-square tests (Table 2) showed significant associations between child’s sex, birth order, residence, division, number of living children, wealth index, maternal education, and household head’s education with stunting, underweight and wasting (p < 0.05). Maternal age at birth was significantly associated with stunting (p = 0.020) and wasting (p = 0.038), but was not significantly associated with underweight (p = 0.789). The child’s age, the sex of the household head and the mother’s age at first marriage were significantly associated with stunting and underweight, but not with wasting. In chi-square test, household size and religion were not significantly associated with any of the malnutrition indicators.

**TABLE 2.**
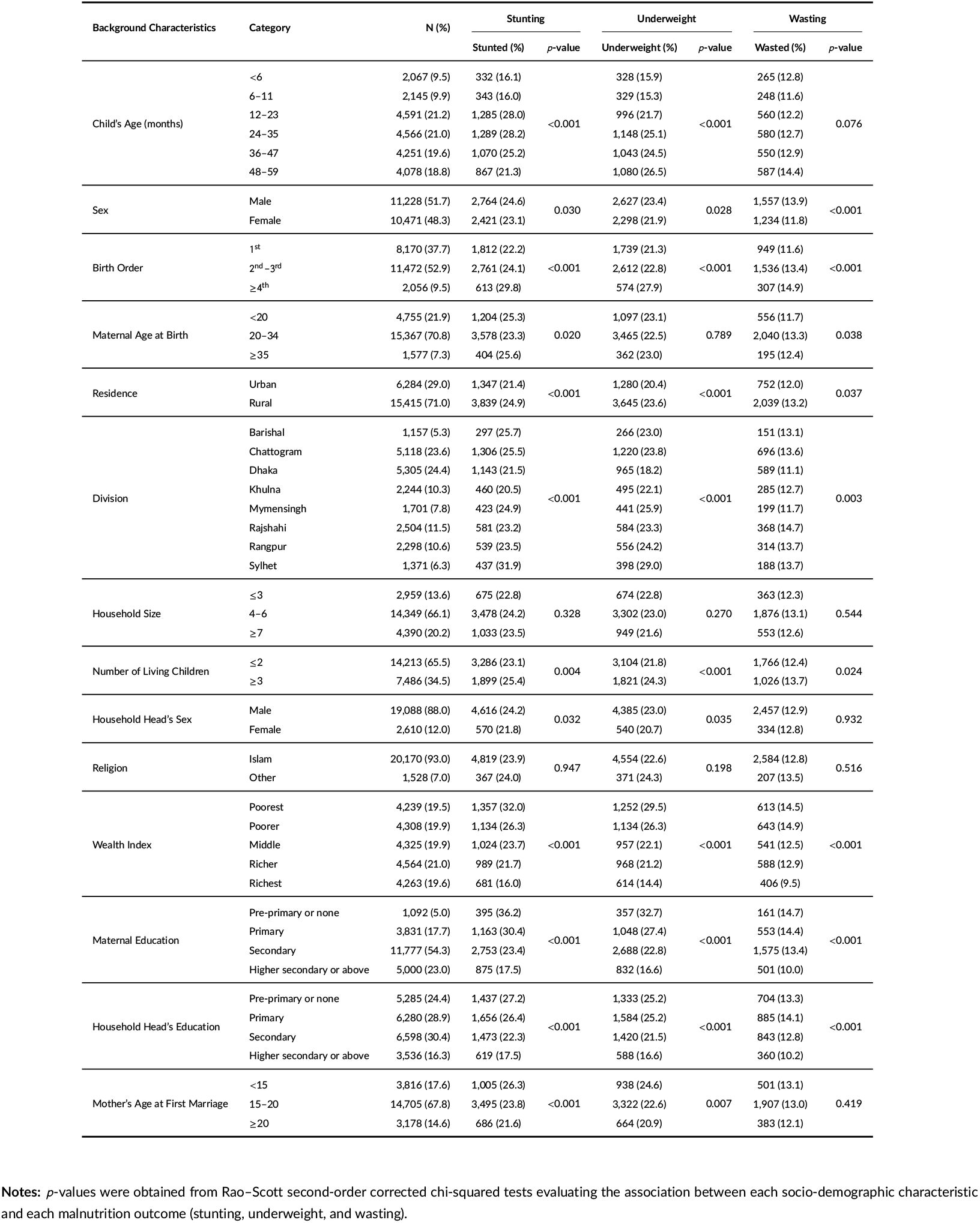
Distribution of child malnutrition types across various socio-demographic factors.

**FIGURE 1.**
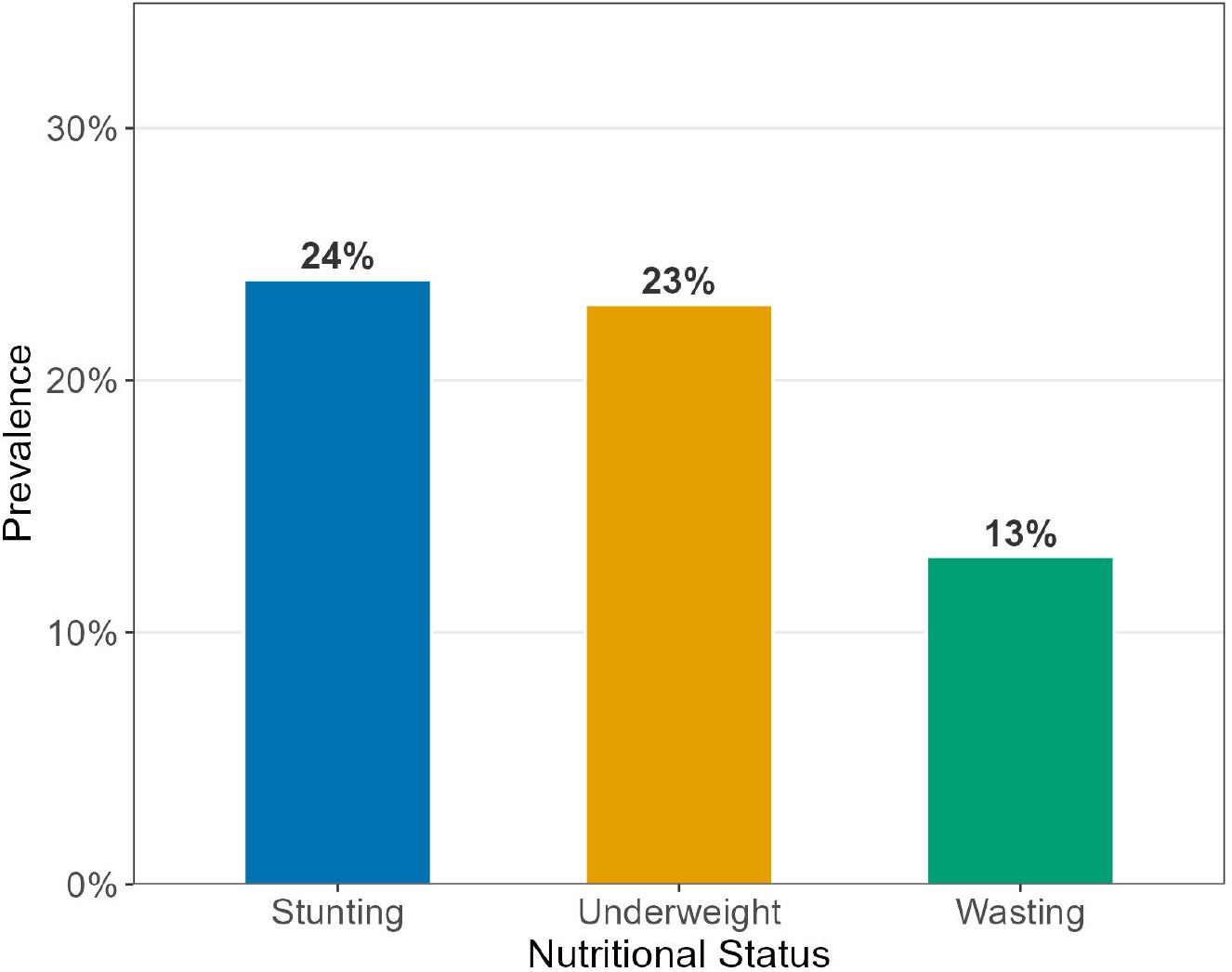
Overall prevalence of child malnutrition indicators among children aged 0–59 months in Bangladesh. Bars represent the weighted prevalence of stunting, underweight, and wasting based on Bangladesh MICS7 data.

Age-related patterns varied somewhat across outcomes. Stunting grew quickly after the first six months of life, increasing from 16.1% in the youngest infants to a high of 28.2% in the 24–35 month age group, and then leveled off somewhat in older children. Conversely, the rise in underweight was more linear, reaching a peak (26.5%) among children nearing their fifth birthday. Across all three outcomes, a persistent sex gap was observed with boys being more affected than girls in every indicator: stunting (24.6% vs. 23.1%), underweight (23.4% vs. 21.9%) and especially wasting (13.9% vs. 11.8%). Malnutrition also closely followed birth order. Fourth or later-born children had the worst outcomes in all categories (29.8% stunted, 27.9% underweight, 14.9% wasted), around 6-8 percentage points higher than first-born children.

All three outcomes were associated with place of residence. Stunting and underweight were more prevalent in rural areas than in urban areas, while a smaller but statistically significant difference was seen for wasting. The highest burden of stunting (31.9%) and underweight (29.0%) was found in Sylhet and the lowest burden of stunting (20.5%) and underweight (18.2%) was shared between Khulna and Dhaka, respectively. The bivariate results showed the most salient pattern of socioeconomic gradients. From poorest to richest wealth quintile, stunting dropped from 32.0% to 16.0%, underweight from 29.5% to 14.4% and wasting from 14.5% to 9.5%. A similar gradient was seen for maternal schooling; children of mothers with no formal education had almost double the prevalence of stunting and underweight as children of mothers who completed higher secondary school or more (36.2% vs. 17.5% and 32.7% vs. 16.6%, respectively). The gradient of household head’s education was similar but slightly flatter.

After adjusting for all covariates in the multivariable survey-weighted logistic regression models, independent associations of several factors with stunting, underweight, and wasting remained, while some bivariate associations were attenuated (Table 3).

**TABLE 3.**
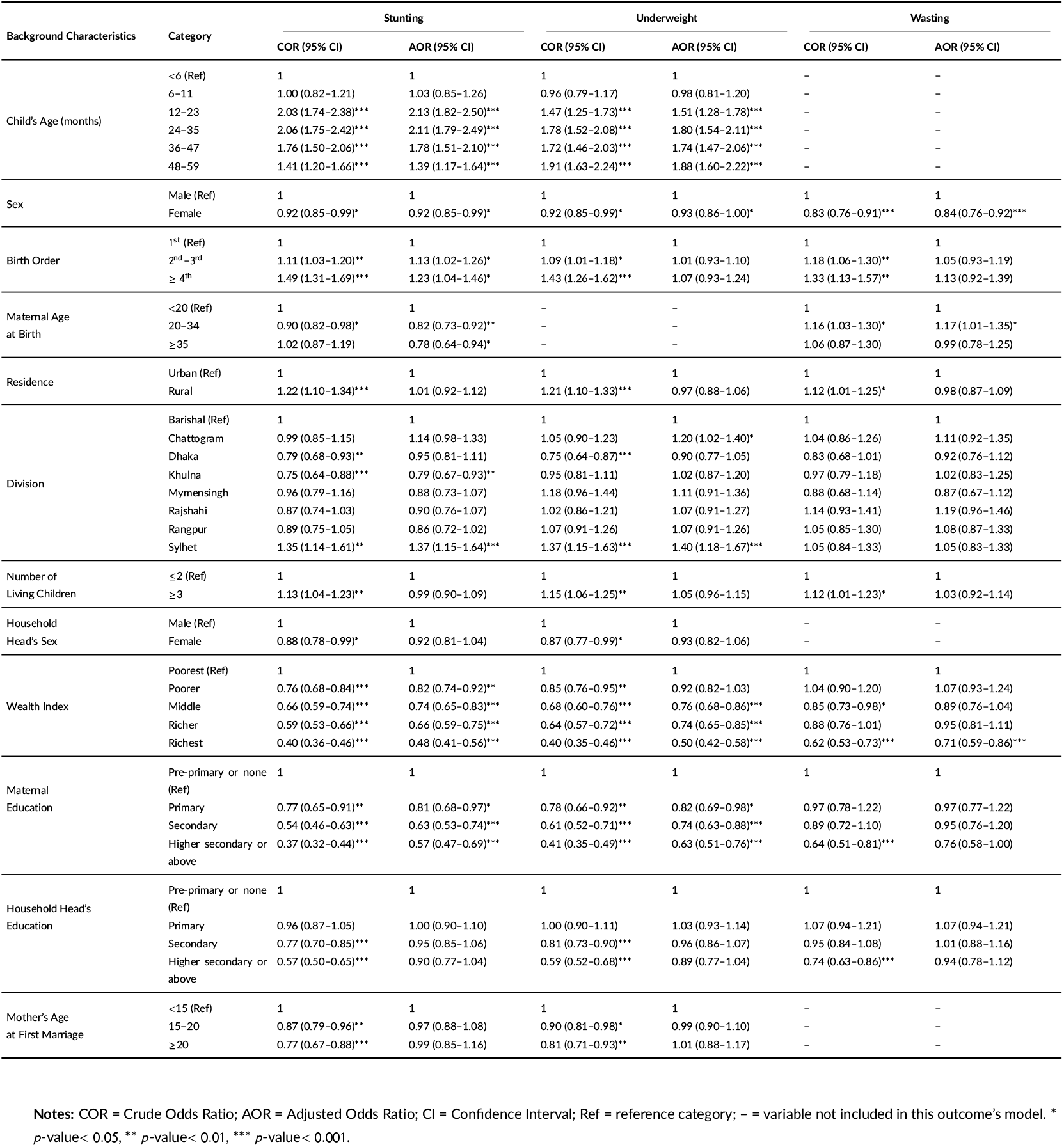
Unadjusted and adjusted odds ratios and 95% confidence intervals for factors associated with stunting, underweight, and wasting among children under five years of age.

Following adjustment, age was still strongly associated with stunting, with the highest odds among children aged 12–23 months (AOR = 2.13, 95% CI: 1.82–2.50) and 24–35 months (AOR = 2.11, 95% CI: 1.79–2.49) compared with children younger than 6 months. Female children had 8% lower odds of stunting compared to male children (AOR=0.92, 95% CI: 0.85–0.99). Higher birth order was still associated with greater odds of stunting (birth order ≥4th: AOR = 1.23, 95% CI: 1.04–1.46). Children born to mothers aged 20 years and above had lower odds of stunting than children born to mothers aged below 20 years (20–34 years: AOR = 0.82, 95% CI: 0.73–0.92; ≥35 years: AOR = 0.78, 95% CI: 0.64–0.94).

Regional disparities persisted after adjustment: children in Sylhet had 37% higher odds of stunting compared to those in Barishal (AOR = 1.37, 95% CI: 1.15–1.64), whereas children in Khulna had 21% lower odds (AOR = 0.79, 95% CI: 0.67–0.93). A strong wealth gradient remained, with children from the richest households having 52% lower odds of stunting than those from the poorest households (AOR = 0.48, 95% CI: 0.41–0.56). Maternal education was also independently protective, with children of mothers with higher secondary education or above having 43% lower odds of stunting than children of uneducated mothers (AOR = 0.57, 95% CI: 0.47–0.69).

The likelihood of underweight increased with child age and was highest among children 48–59 months of age (AOR = 1.88, 95% CI: 1.60–2.22) compared to children under 6 months. Females were less likely to be underweight than males (AOR = 0.93, 95% CI: 0.86–1.00). Children from Chattogram (AOR=1.20, 95% CI: 1.02-1.40) and Sylhet (AOR=1.40, 95% CI: 1.18-1.67) had significantly higher odds of being underweight compared to children from Barishal. Like stunting, the wealth gradient was observed for underweight with children from the richest households having 50% lower odds of being underweight than children from the poorest households (AOR = 0.50, 95% CI: 0.42–0.58). Maternal education independently remained protective, with the lowest odds among children of mothers with higher secondary or above education (AOR = 0.63, 95% CI: 0.51–0.76).

Age was excluded from the wasting model as the bivariate association was not statistically significant unlike stunting and underweight. The odds of being wasted were 16% lower among female children compared to male children (AOR = 0.84, 95% CI: 0.76–0.92). Children born to mothers aged 20–34 years had higher risk of being wasted than children of mothers aged <20 years (AOR = 1.17, 95% CI: 1.01–1.35). No significant association was found between division and wasting after adjustment. Wealth index remained a significant predictor with children from the richest households having 29% lower odds of wasting compared to the poorest households (AOR = 0.71, 95% CI: 0.59–0.86); no other wealth categories reached statistical significance after adjustment. In the adjusted model, wasting was not significantly associated with maternal education, education of household head, birth order, residence, and number of living children.

## 4 DISCUSSION

The findings of this study show that malnutrition remains widespread among children under five, with 24% experiencing stunting, 23% being underweight, and 13% experiencing wasting. The results also reveal that different demographic and socioeconomic characteristics are associated with children’s nutritional status in distinct ways. Stunting increased markedly after infancy, with the greatest vulnerability observed among children aged 12–35 months. One possible explanation is that biological and socioeconomic factors play an increasingly important role in stunting as children grow older [36]. The prevalence of underweight increased with age, with older children more frequently affected than younger children [37]. This age-specific pattern highlights the importance of early childhood as a critical period for the development of stunting and it is consistent with evidence from previous studies [38, 39]. The difference in age patterns exists because stunting measures chronic, structural height deficits that primarily develop and stabilize within the first 1,000 days of life [40]. In contrast, underweight is a composite indicator reflecting both height and immediate weight deficits; as children grow older, it continues to rise due to increasing metabolic demands, cumulative nutritional gaps, and shifts in household feeding priority toward younger siblings [41].

Child sex remained significantly associated with all three forms of malnutrition after adjustment, with female children having lower odds of stunting, underweight, and wasting compared with male children. This consistent pattern suggests that boys may be more vulnerable to childhood malnutrition across different nutritional indicators. Similar sex differences have been reported in previous studies, with evidence showing a higher prevalence of malnutrition among boys than girls [42, 43, 44]. This higher burden among boys may partly reflect their greater biological susceptibility to morbidity compared with girls [45].

Higher birth order was significantly associated with an increased likelihood of stunting. It has also been found in previous studies that higher birth order is related to a greater prevalence of stunting [46, 47]. Probably, mothers with many children have overall less time to extend care equally to each of the children than those who have fewer children [48].

Maternal age at birth was significantly associated with stunting. Children born to mothers aged 20 years or older had lower odds of stunting than children born to mothers younger than 20 years [49]. Biologically, adolescent mothers are still growing, so their own nutritional needs compete with those of the developing foetus. This raises the risk of intrauterine growth restriction, low birth weight and preterm birth, all of which predispose a child to poor linear growth [50]. Behaviourally, younger mothers may breastfeed for a shorter duration and may be less experienced in complementary feeding, hygiene and care practices [51]. Socially, they tend to have less education, lower socioeconomic status, less autonomy in household decisions and more psychological stress from limited resources [51]. Together, these factors may limit the quality of care and feeding a child receives, increasing exposure to inadequate dietary intake and infection, which over time may impair linear growth.

Regional disparities in childhood malnutrition remain stark after adjustment. Children living in the Sylhet division face significantly higher odds of both stunting and underweight compared to those in other regions [52, 19, 53]. This finding points to a well-documented regional paradox in Bangladesh: despite having relatively lower poverty rates [54]and better overall food security [55], Sylhet consistently lags behind in child health outcomes [36]. This trend is heavily driven by a severe educational deficit. Sylhet suffers from the lowest secondary school completion rates in the country. Because mothers are typically a child’s primary caregiver, this lack of formal education directly limits access to crucial health information, proper hygiene practices, and nutritional literacy [36]. Furthermore, Sylhet’s unique geography creates severe infrastructural barriers because the region is characterized by remote tea gardens and vast wetlands known as haors that flood during the monsoon season. This difficult terrain heavily restricts local transport, which hinders children from attending school and physically cuts off vulnerable families from healthcare clinics and food markets [36].

Place of residence was associated with all three nutritional outcomes in bivariate analysis, with rural children showing a consistently higher burden of stunting, underweight, and wasting than urban children, though the urban-rural gap was narrower for wasting than for the other two outcomes. This bivariate disparity is consistent with observations from other developing-country contexts, where the rural–urban gap in child malnutrition has historically persisted due to ongoing economic hardship, limited access to health services, and gaps in parental education [56, 57]. However, this association did not remain significant once wealth, maternal education, and division were accounted for in the multivariable models, suggesting that the crude rural-urban disparity largely reflects underlying differences in these factors between rural and urban populations, rather than residence itself exerting an independent effect [56].

The likelihood of undernourishment consistently decreased as maternal education level increased, with children of mothers who completed higher secondary education or above facing the lowest odds of stunting and underweight [58, 52]. Educated mothers typically possess a greater understanding of preventative health and sanitation practices, which empowers them to provide higher-quality care for their offspring [59, 60, 61]. Additionally higher maternal education is associated with a higher likelihood of exclusive breastfeeding during the first six months of a newborn’s life and the timely initiation of complementary foods [62]. Furthermore, educated mothers are more likely to make better use of health care services for their children, make efficient use of household family resources, and demonstrate a greater willingness to utilize family planning [63, 64]. Finally, more educated mothers may earn higher income, providing them with more opportunity to directly invest in the health, nutrition, and hygiene requirements of their children [65].

Household wealth showed a strong socioeconomic gradient in childhood malnutrition, with children from wealthier households consistently less likely to experience stunting and underweight [19]. The lower odds of child stunting associated with a higher wealth index might be explained by the fact that children from wealthier families have better access to health facilities, better environmental conditions such as potable water and sanitation, and access to sufficient food [36]. Furthermore, parents from the wealthiest families often have a higher education and may be more responsible for the health of their children when compared with parents from the poorest households [36]. Other studies have shown that low quality housing and a lack of access to potable water are significantly associated with child stunting [66]. Conversely, in our adjusted model, this protective socioeconomic gradient was much less consistent for childhood wasting, as only children from the richest households exhibited significantly lower odds of being wasted. This specific discrepancy highlight that household asset ownership may not uniformly protect children from acute wasting. According to Fernandez et al., wasting reflects an immediate weight deficit relative to height resulting from short term nutritional or infectious insults, which differs from the linear growth retardation that characterizes chronic stunting [67]. Therefore, while intermediate wealth levels may provide enough stability to prevent chronic stunting over time, they may not offer sufficient economic cushioning to shield children from the sudden biological and environmental shocks that trigger acute wasting.

## 5 CONCLUSION

Childhood malnutrition remains a major public health concern in Bangladesh. This study found that children’s nutritional status was associated with several demographic and socioeconomic factors. Boys, children from poorer households, and children whose mothers had lower levels of education were particularly vulnerable to malnutrition. Child age, birth order, maternal age at birth, and geographic division were also associated with specific forms of malnutrition. Regional disparities were evident, with children in Sylhet experiencing a particularly high burden of stunting and underweight.

Reducing childhood malnutrition requires coordinated efforts from the government, non-governmental organizations, healthcare providers, and local communities. Nutrition programs should prioritize socioeconomically disadvantaged families, strengthen maternal education and nutrition awareness, and provide appropriate nutritional support during the most vulnerable stages of childhood. Region-specific interventions, particularly in areas with a high burden of malnutrition, should also be considered. Continued nutritional surveillance is essential for identifying vulnerable groups, assessing the effectiveness of existing programs, and supporting Bangladesh’s progress toward national and global nutrition targets.

## 6 LIMITAION & FURTHER ANALYSIS

This study has limitations. First the cross-sectional design prevents causal inference so the associations identified should not be interpreted as causal. Second determinants such as dietary intake, breastfeeding, birth weight, recent illness and water, sanitation and hygiene (WASH) conditions were not included in the analysis so residual confounding is possible. Third the exclusion of children, with implausible Z-scores or missing covariate data may have introduced selection bias. Nevertheless the large representative sample and survey-weighted analysis support the generalizability of the findings.

## Data Availability

This study used publicly available data from the Bangladesh Multiple Indicator Cluster Survey (MICS), conducted
by the Bangladesh Bureau of Statistics with support from UNICEF. The dataset can be accessed at https://mics.unicef.org/

## Abbreviations

AOR: adjusted odds ratio
BBS: Bangladesh Bureau of Statistics
BDHS: Bangladesh Demographic and Health Survey
CI: confidence interval
COR: crude odds ratio
EA: enumeration area
HAZ: height-for-age Z-score
MICS: Multiple Indicator Cluster Survey
PPS: probability proportional to size
Ref: reference category
SD: standard deviation
SDG: Sustainable Development Goal
UNICEF: United Nations Children’s Fund
WAZ: weight-for-age Z-score
WHO: World Health Organization
WHZ: weight-for-height/length Z-score.

## Acknowledgements

The authors are grateful to the Bangladesh Bureau of Statistics and UNICEF for making the nationally representative Multiple Indicator Cluster Survey (MICS) data available for this study.

## Author Contributions

YAO and KK contributed equally to the study design, statistical analysis, and manuscript drafting. YAO and KK revised the manuscript. All authors read and approved the final manuscript.

## Availability of data and materials

This study used publicly available data from the Bangladesh Multiple Indicator Cluster Survey (MICS), conducted by the Bangladesh Bureau of Statistics with support from UNICEF. The dataset can be accessed at https://mics.unicef.org/.

## Ethics Statement

This study is a secondary analysis of data from the Multiple Indicator Cluster Survey (MICS), implemented by the Bangladesh Bureau of Statistics with support from UNICEF. The survey received ethical clearance from the relevant national ethics review bodies, and informed consent was obtained from all respondents during data collection. Because the present analysis used anonymized data, no further ethical approval was required.

